# Single-strand RPA for rapid and sensitive detection of SARS-CoV-2 RNA

**DOI:** 10.1101/2020.08.17.20177006

**Authors:** Youngeun Kim, Adam B. Yaseen, Jocelyn Y. Kishi, Fan Hong, Sinem K. Saka, Kuanwei Sheng, Nikhil Gopalkrishnan, Thomas E. Schaus, Peng Yin

## Abstract

We report the single-strand Recombinase Polymerase Amplification (ssRPA) method, which merges the fast, isothermal amplification of RPA with subsequent rapid conversion of the double-strand DNA amplicon to single strands, and hence enables facile hybridization-based, high-specificity readout. We demonstrate the utility of ssRPA for sensitive and rapid (4 copies per 50 µL reaction within 10 min, or 8 copies within 8 min) visual detection of SARS-CoV-2 RNA spiked samples, as well as clinical saliva and nasopharyngeal swabs in VTM or water, on lateral flow devices. The ssRPA method promises rapid, sensitive, and accessible RNA detection to facilitate mass testing in the COVID-19 pandemic.

## Introduction

Effective and accessible mass testing can help to limit the spread of the current SARS-CoV-2 pandemic. While serology testing reveals recent and past exposure, RNA testing allows early detection of active infection. Standard RT-qPCR^1^ and relatives^2–10^ achieve high analytical sensitivity (1-100 copies of viral RNA per input μl)^11^, but take hours and require relatively complex equipment. New sequencing approaches promise high multiplexing but slow response further^12–15^. In contrast, isothermal methods such as Recombinase Polymerase Amplification (RPA)^16–22^, Recombinase-Aided Amplification (RAA)^23^, Nicking Enzyme Amplification Reaction (NEAR)^24^, Loop-mediated isothermal Amplification (LAMP)^25–39^, and Rapid Amplification (RAMP, an RPA + LAMP combination)^25^ typically detect 10-1000 copies of RNA per reaction with minimal or no instrumentation (see review^40^). The RPA reaction can generate double-stranded DNA (dsDNA) amplicons particularly quickly, but its recombinase-driven priming process is prone to multi-base mismatching that necessitates an additional specificity check^39,41^. Augmentation of RPA with conditionally extensible primers or cleavable inter-primer probes enhances specificity^16,42,43^, but tends to reduce reaction speed. Alternatively, Cas12^34^ or Cas13^19,44^ nucleases applied to amplification products generate signal in a sequence specific manner, but incur substantial increases in workflow complexity and reaction time. Here, we describe “single-strand RPA” (ssRPA) method, which applies (1) rapid amplification of dsDNA, (2) conversion to ssDNA, and (3) sequence-specific, hybridization-based readout, arranged to maintain both optimal speed and accuracy (Fig. 1a). For amplification, we apply basic RT-RPA^16^ in the absence of any specificity-enhancing components that may inhibit speed. For ssDNA conversion, we use an exonuclease to digest strands except those chemically protected. Finally, hybridization-based readout is shown with LFDs.

**Figure 1:**
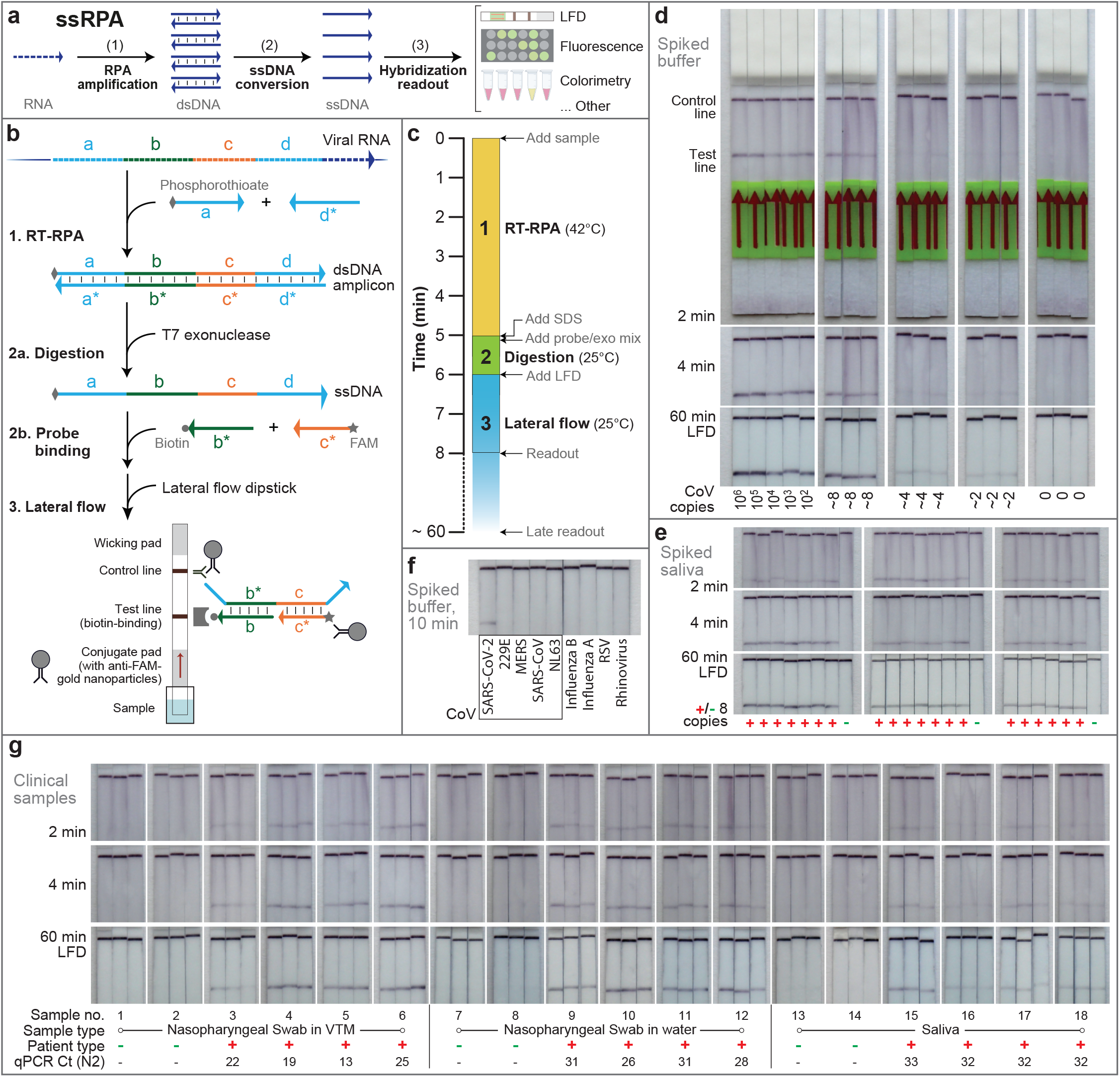
ssRPA assay design, workflow, and characterization. **(a)** Key to ssRPA design is the rapid generation of many ssDNA copies from a single RNA target. ssDNA output offers straightforward specific readout by fluorescence or colorimetric/visual methods such as lateral flow devices. **(b)** Step 1: Target viral RNA region (of domains *a-b-c-d*) is reverse transcribed into cDNA via extension of the reverse primer (*d\**) by the Reverse Transcriptase in the reaction mixture^48^. Subsequently, the cDNA is amplified via isothermal RPA at 42 °C by templated extension of the forward (*a*) and reverse primers (*d\**). The forward primer has a 6-nucleotide long poly-T segment with phosphorothioate bonds. Step 2a: Products of RPA are amended with SDS and transferred to the exo/LFD buffer that contains T7 exonuclease (a dsDNA-specific 5′ to 3′ exonuclease) and detection probes. The resulting mixture is incubated for 1 min at ambient temperature and reverse strand of the dsDNA amplicon products get preferentially digested yielding ssDNA amplicon (*a-b-c-d*) homologous to the target RNA sequence. Step 2b: The 3′ biotin (*b\**) and 5′ FAM (*c\**) modified detection probes, already added, hybridize to target sequences. These probes make the assay directly compatible with commercially available test strips that feature a biotin-binding test line and gold nanoparticles conjugated to rabbit anti-FAM IgG on the conjugate pad. Step 3: The test strip is vertically inserted into the resulting 100 µl mixture. The right ssDNA amplicon acts as a bridge that binds both the biotin-probe and the FAM-probe independently resulting in immobilization of the complex at the test line, where formation of a colored line indicates a positive result. The control line formed of rabbit secondary antibodies captures the remaining gold nanoparticle conjugates by binding to rabbit anti-FAM IgG. **(c)** Timeline of the assay shows the incubation conditions and duration of the 3 main steps in ssRPA: (1) RT-RPA, (2) exonuclease digestion and (3) lateral flow. The test line and control line can be visualized as early as 1-2 min or as late as 60^+^ min without false positives. **(d)** Sensitivity of ssRPA-LFD demonstrated by serial dilution of synthetic SARS-CoV-2 full genome standard (Twist) from ∼ 1,000,000 copies down to ∼ 2 copies per reaction. A 5 µl volume of genomic viral RNA in DNase/RNase-free water was used as input for a 50 µl reaction volume. After RT-RPA for 5 minutes at 42 °C, 8 µl product was mixed with 12 µl of 10% SDS and then transferred into 80 µl of exo/LFD buffer. Following 1 min T7 exonuclease digestion at room temperature, samples were applied to commercial HybriDetect strips for ≥ 1 min. A time series for the same strip is shown in each column. Note the zero test line signal in negative controls, even at 60 min. **(e)** Using the same procedure as in (d), but spiking 8 copies cultured, heat inactivated SARS-CoV-2 virus (BEI) into 5 µl of SARS-CoV-2-negative human saliva in 20 repeats. No-template negative controls are also shown. (**f**) Specificity was shown by testing 8 other respiratory virus genomic samples, including genomically similar coronaviruses 229E, MERS, (2003) SARS-CoV, and NL63, and alternative diagnoses of influenza B, influenza A, respiratory syncytial virus (RSV), and rhinovirus 17, each at >10^5^ copies per assay and spiked in DNase/RNase-free water. A sample with virus-derived SARS-CoV-2 RNA at low copy is shown as a positive control. Strips show the readout at 10 min of lateral flow. **(g)** Clinical samples were taken as NP swabs in VTM, NP swabs in water, or saliva, from SARS-CoV-2 positive and negative patients alike, and heat inactivated at vendor for safety. Sample order was randomized prior to ssRPA, and technician was blinded as to patient type or qPCR status to avoid experimental bias. During assay, samples further underwent a 2 min extraction protocol with Lucigen DNA extraction buffer at 50% dilution and 95 °C, and were tested at 10% v/v into ssRPA for 5′ spike targets (see Methods). Non-extracted samples yielded little or no signal (Fig. S6). Presumptive diagnosis from supplier testing is noted. Concomitant qPCR quantification in our lab, using CDC N2 primers from buffer-exchanged samples, is shown with each result (See also Fig. S7).

## Results

We selected the 5′ end of the SARS-CoV-2 spike protein sequence as our main detection target. Per the detailed protocol of Fig. 1b (and Supplementary Protocol), we diluted the sample in a basic RT-RPA reaction mixture, modifying the forward primer with a 5′ tail of 6 phosphorothioate-linked bases to confer exonuclease protection^45,46^. The reaction was run for 5 min at 42°C on a heating block. A sample of the product was then treated with sodium dodecyl sulfate (SDS) and diluted into an exonuclease and lateral flow (exo/LFD) buffer, where the unprotected strand in the dsDNA was rapidly (≤ 1 min) digested by a T7 exonuclease to yield the protected ssDNA target^45,46^. A pair of 3′-biotin and 5′-FAM modified probes in the digestion buffer were available to target sequences in between the amplification priming domains, providing specificity not achievable with RPA priming alone. The correct target ssDNA therefore acted as a bridge that co-localized both detection probes within an LFD, ultimately binding gold nanoparticles to the biotin-ligand test line to produce visual readout as early as 1-2 minutes. Full amplification and detection timelines are described in Fig. 1c.

We first tested ssRPA on buffer-spiked samples. Figure 1d (and Supplementary Fig. S1) shows LFD detection of synthetic SARS-CoV-2 full genome standard serially diluted in DNase/RNase-free water, photographed at multiple intervals on the same strips. Concentrations of input RNA were quantified by RT-qPCR and direct comparison to spectrophotometrically-quantified synthetic standards (Fig. S2). Results show detection of 8 copies per 50 µL reaction by naked eye within 8 min (2 min LFD), or 4 copies within 10 min (4 min LFD), and a dynamic range of at least 5 orders of magnitude. No test line signal formed for the no-template negative controls over >60 min of LFD incubation. To test for assay reliability, we spiked 5 µL human saliva with 8 copies of cultured, heat-inactivated virus and showed 20 of 20 positive tests within 8 min (Figs. 1e and S3). To test for specificity, we performed ssRPA on water spiked with viral RNA from 8 other respiratory viruses, including genomically similar coronaviruses and alternative diagnoses, each at >10^5^ copies per assay. None exhibited false positive signals (Figs. 1f and S4). Finally, we performed a blinded assay on clinical samples, with a technician unaware of patient type or qPCR result correctly identifying the type (positive or negative) of all 18 samples at 2 min LFD time. Nasopharyngeal (NP) swabs stored in viral transport media (VTM), NP swabs stored in water, and saliva samples were processed by single-tube RNA extraction (1:1 mixture with extraction buffer, 95 °C × 2 min)^47^ and used at 10% v/v in RT-RPA (Figs. 1g and S5). All clinical samples were also semi-quantified by RT-qPCR, using column extraction/concentration and CDC N2 primers (Fig. 1g, S7). Results of this limited study demonstrate 100% sensitivity and 100% specificity across all sample types, including NP swabs in water and saliva samples with lower titers (high C_t_).

## Discussion

The ssRPA method combines the speed of established RT-RPA^16^ with the sequence specificity of ssDNA hybridization by serially applying RPA and exonuclease steps. It enables best-in-class performance on the principal axes of reaction time and sensitivity, enabling the detection of 8 copies in 8 min or 4 copies in 10 min. The conceptual framework could be generalized to other isothermal readout methods with dsDNA output for achieving optimal sensitivity and speed. The present method may also be further developed to achieve single-nucleotide specificity, for example by using toehold probe readout^19^, and spatial multiplexing readout on LFDs with multiple test positions. We also expect future development, including simplified workflow and use of lyophilized reagents for ambient distribution and storage, will further facilitate mass testing.

## Data Availability

All relevant data are included within the manuscript and supplemental information.  Please contact corresponding authors for more information.

## Acknowledgements

We acknowledge support from NIH 1DP1GM133052-01 and the Wyss Institute Molecular Robotics Initiative. We thank Girija Goyal and Don Ingber for inactivated SARS-CoV-2 samples, and Robert Rasmussen for help with safety and regulatory compliance. Noted reagents were obtained through BEI Resources, NIAID, NIH.

## Supplemental Information

### Methods

#### Sample sources

Synthetic SARS-CoV-2 RNA (Twist Biosciences, 102019) was used in sample qPCR quantification and in the serial dilution experiment of Fig. 1d. Synthetic fragment RNA (IDT), constituting the 76 nt region beginning at the 5′ end of the spike sequence (“5′ S region”), was used in sample qPCR quantification and ordered as Ultramers at 20 nmol scale. All synthetic primers and probes (IDT) were chemically synthesized with any specified modifications, ordered at 100 or 250 nmol scale, desalted or PAGE-purified, and used as is. Viral genomic RNA (isolated from infected cells) from coronaviruses 229E (ATCC, VR-740D), MERS (BEI, NR-50549), SARS-CoV-1 (BEI, NR-52346), and NL63 (BEI, NR-44105), as well as influenza A (ATCC, VR-1736D), influenza B (ATCC, VR-1535D), respiratory syncytial virus (ATCC, VR-1580DQ), and rhinovirus (ATCC, VR-1663D), were used for the specificity experiment in Fig. 1e. Cultured and heat-inactivated SARS-CoV-2 viral RNA (BEI, NR-52286) was used in specificity (Fig. 1e) and in spiked saliva (Fig. 1f) experiments. Pooled human saliva from ≥ 3 de-identified donors (Lee Biosolutions, 991-05-P) was collected prior to November 2019 and used to prepare the contrived samples. All clinical samples (Fig. 1g) were purchased from BioCollections Worldwide, Inc. and heat-inactivated at 95 °C for 5 min prior to shipping.

#### Primer and probe design

Relatively clean RPA products are the result of multiple primer pairs designed *in silico* for the appropriate salt and temperature conditions (NUPACK.org) and extensively tested empirically, with the goal of minimizing primer dimers and other unintended reactions that might slow the targeted amplification. RPA primer sequences and LFD probes for SARS-CoV-2 5′ spike target were as follows, where “*” denotes a phosphorothioate bond (for exonuclease protection), “/iFluorT/” denotes an internal FAM fluorophore (for nanoparticle capture), “/3ddC/” denotes 3′ dideoxycytidine (to prevent 3′ end extension), and “/3Bio/” denotes 3′ biotin (for test line capture). Here, the probe sequences partly overlap with primer sequences, but not enough to bind significantly to those sequences in isolation. This overlap is not necessary. Also note that, for extended protection, an internal fluorescein with poly-phosphorothioate protection is noted here, though we have had similar results with a simple 5′ /56-FAM/ moiety. Probes and primers for 5′ spike were: Fwd primer: T*T*T*T*T*T*TAACATCACTAGGTTTCAAACTTTACTTGC; Rev primer: CCAACCTGAAGAAGAATCA CCAGGAGTCAA; FAM LFD probe: T*T*T*T*T*T*T /iFluorT/ TTTTTTTTTTTTT AGGAGTCAA ATAACTT/3ddC/; Biotin LFD probe: T*T*T*T*T*T* TATGTAAA GCAAGTAAAG TTTTTTTTTTTTTTT /3Bio/.

#### Sample preparation

SARS-CoV-2 samples were quantified in-house, comparing fluorometrically-quantified synthetic RNA fragments (IDT), Twist Biosciences qPCR RNA standards, and culture-derived BEI viral genome samples, all diluted in DNase/RNase-free water (See SI Fig. S2). Synthetic fragment (IDT) RNA segment was quantified by Nanodrop spectrophotometry by blanking with IDTE buffer (IDT) and then measuring absorbance at 260 nm with a 2 µl sample volume. Ten-fold serial dilutions of the synthetic fragment (IDT) RNA segment were made from 10^7^ copies/µl down to 10^3^ copies/µl in DNase/RNase-free water, using low binding tips and tubes to avoid sample loss. Similarly, Twist synthetic SARS-CoV-2 RNA were serially diluted in ten-fold (0.1×, 0.01×, 0.001×) for comparison. Amplification by qPCR (Bio-Rad, CFX Connect) was then performed with 4× TaqPath 1-Step RT-qPCR master mix (Life Science, A15300), 5′ spike primers (100 nM), and EvaGreen dye (Thermofisher) at 20 µl total volume with 1 µl sample volume. A linear regression was performed on the Ct and expected dilution concentrations of the standard (R^2^ = 0.99745), and in turn used to convert sample Ct to absolute quantities of 552000, 55400, and 7200 copies/µl, respectively. This quantified Twist sample was then used in a dilution series to quantify the culture-derived (BEI) RNA using Charite “E” primer and sequence-specific probe set (IDT). Here, qPCR was performed with 5 µl of 4× TaqPath 1-Step RT-qPCR master mix, 3 µl of Charite “E” primers/probes, and 7 µl of DNase/RNase-free water at 20 µl total volume with 5 µl input sample volume. Simple and contrived samples were prepared by further diluting quantified samples as necessary, spiking into DNase/RNase-free water or pooled human saliva at ∼ 2 or more copies per 5 µl, and used directly in RT-RPA reactions. For the specificity experiments, genomic RNA from 8 respiratory viruses were spiked in DNase/RNase-free water at 10^5^ copies/µl, unless noted (in which case quantification was not supplied from source). As a positive control, heat inactivated SARS-CoV-2 virus was used at 100 copies/µl. All were diluted 1:50 (1 µl input into 50 µl total reaction volume) in the RT-RPA reaction mixture.

#### RT-RPA

A mixture of 2.5 µl each of 10 µM forward and reverse primers to the specified target, 29.5 µl of TwistAmp Basic RPA rehydration buffer (TwistDx, TABAS03KIT), 5 µl of DNase/RNase-free water, and 0.5 µl of Protoscript II reverse transcriptase (NEB, M0368S) were vortexed briefly and added to the TwistAmp lyophilized powder mix. Then, 5 µl of sample was added to the tube while 5 µl of 280 mM Magnesium Acetate was added to the reaction tube lid. The 50 µl mixture was vortexed briefly, spun down, vortexed again, spun down once more and immediately incubated at 42 °C for 5 min in a standard PCR machine (Applied Biosystems, 4484073) or heating block (Benchmark Scientific, BSH300).

#### Amplicon digestion

During amplification, a digestion and LFD buffer mixture was prepared with 64 µl of LFD running buffer (Milenia, MGHD 1), 1 µl of 10 µM biotin probes, 1 µl of 10 µM FAM probes, 10 µl of 10× NEBuffer 4 (NEB, B7004S), and 4 µl of T7 exonuclease (NEB, M0263S), in a total volume of 80 µl. The mixture was vortexed briefly and added to a 2 mL lo-bind tube (Eppendorf). Once complete, 8 µl of the RT-RPA reaction was subsequently mixed thoroughly and completely with a 10% Sodium Dodecyl Sulfate (SDS) at a ratio of 8 µl sample:12 µl SDS to inactivate enzymes. This 20 µl mixture was then added to the 80 µl digestion mixture above and incubated for 1 min at room temperature.

#### Electrophoresis

All gels (8 × 8 cm) were denaturing PAGE at 15% polyacrylamide (Invitrogen, EC6885BOX), run in 1× TBE buffer that was diluted from 10× TBE (Promega, V4251) with filtered water, at 65 °C, 200V, for 30 min. Immediately after 5 min of RT-RPA, 5 µl of products were directly mixed with 5 µl of formamide dye consisted of 100% formamide (Thermofisher) and Bromophenol blue dye (Sigma Aldrich). Gels were then removed from cassettes, stained in 1× SybrGold (Life Technologies) for 3 min, and imaged with a Typhoon scanner (General Electric). Ladders shown are 25–766 nt DNA (NEB, #B7025).

#### Lateral flow assay

A standard HybriDetect LFD strip (Milenia Biotec, MGHD 1) was inserted into the 2 mL Eppendorf tube above, arrows pointing up/away from the mixture, with care taken not to handle the strip roughly. Note that strips are covered with a membrane that protects the nitrocellulose and supported by a semi-rigid backing card. The strip was incubated for 2 min or longer, as desired.

### Protocol: ssRPA-LFD

#### Materials needed

- TwistAmp Basic Kit (thawed at ambient temperature); this protocol is specifically written to use these dried powder kits (Basic Kit) and has yet to be verified for the TwistAmp Liquid Kit
- Forward primer 10 µM (IDT)
- Reverse primer 10 µM (IDT)
- RNA template
- Protoscript II reverse transcriptase (NEB, M0368S)
- DNase/RNase-free water
- SDS
- Lateral flow strips and buffer (Milenia, MGHD 1)

**Quick RNA extraction protocol** (adapted from Ladha et al, doi: 10.1101/2020.05.07.20055947)

0.1. Take 5 µl of patient sample (whether VTM, water, or saliva)
0.2. Mix with 5 µl of Lucigen DNA extraction buffer
0.3. Incubate at 95 °C for 2 minutes
0.4. Take out the tube and immediately keep on ice
0.5. Use 5 µl (out of the total 10 µl per sample) for ssRPA

#### ssRPA protocol

##### Step 1: Set up and run RPA (on PRE-AMPLIFICATION BENCH)

* Please have the heat block set up before setting up reaction to ensure reaction time.

1.1. Prepare (per reaction) in the following order at room temperature:

|  |  |
| --- | --- |
| 5 μl | DNase/RNase-free water |
| 29.5 μl | Rehydration buffer (included in TwistDX kit) |
| 2.5 μl | 10 μM forward primer |
| 2.5 μl | 10 μM reverse primer |
| 0.5 μl | Protoscript II Reverse Transcriptase |

Vortex ∼ 3 seconds and spin briefly (∼ 3 seconds).

If making a master mix, ensure that ∼(n+1)× master mix solution is mixed for n samples to ensure that all samples get the full, correct amount of master mix.

1.2. Add above reaction to a TwistAmp Basic reaction (dried powder included in TwistDX kit). Do not mix or pipette up and down. Then add 5 µL of RNA template directly into the tube (or water for the negative control).

1.3. Add 5 µL of 280 mM Magnesium Acetate (included in TwistDX kit) to tube lid (this way, MgOAc is kept separate in the tube lid prior to overall mixing). Carefully close tube lid without touching the inside of the tube lid (to avoid any contamination from previous amplicons or other sources), vortex briefly (∼ 2 seconds), spin down briefly (∼ 2 seconds on a micro-centrifuge), then vortex again ∼ 2 seconds to start reaction. Spin briefly (∼ 2 seconds) before the next step.

1.4. Immediately, incubate it at 42 °C for 5 minutes.*

##### Step 2: Set up LFD (on POST-AMPLIFICATION bench)

2.1. While RPA is running, prepare (per reaction) the following in a 2 mL lo-bind tube:

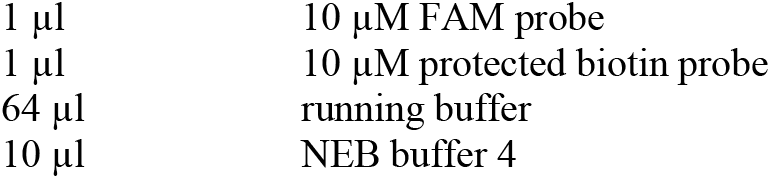

Vortex and spin briefly, then to each add:

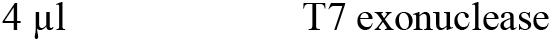

Vortex and spin briefly.

2.2. Add 12 µL of 10% SDS to lid of the tube. (Avoid long spin-down of 10% SDS solution, and make sure to store at room temperature prior to this addition.) Once RPA reaction is complete, immediately vortex RPA samples for 3-5 seconds. Then add 8 µL of RPA sample to lid and pipette up and down 25-30 times quickly and vigorously until white bubbles appear to mix RPA sample with SDS. Immediately, spin solution down, vortex for 5 seconds, spin for 5 seconds.

2.3. Incubate at room temperature for 1 minute (for exonuclease digestion).

2.4. Place the lateral flow strip into tube, incubate for 2 minutes (but may wait up to an hour).

**Figure S1:**
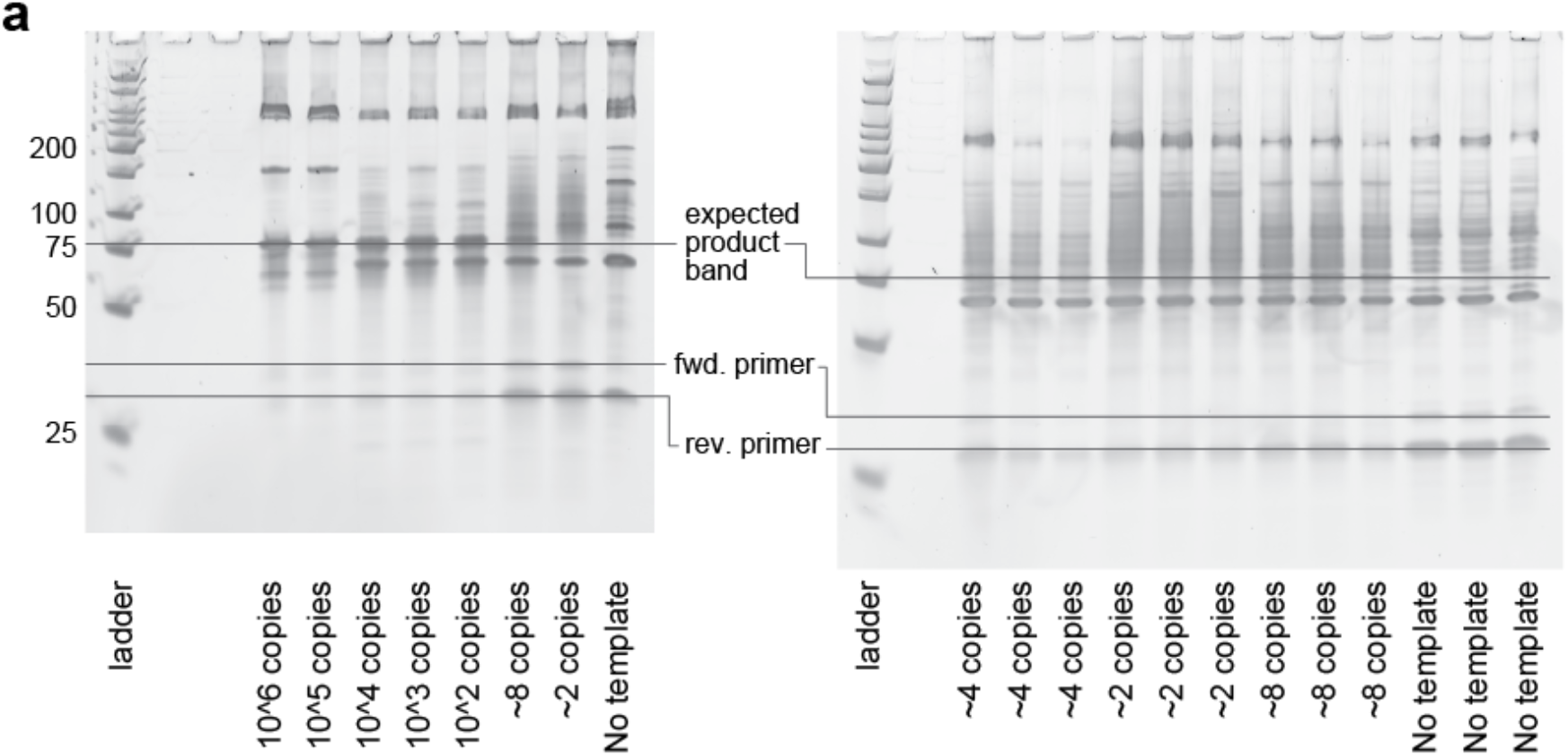
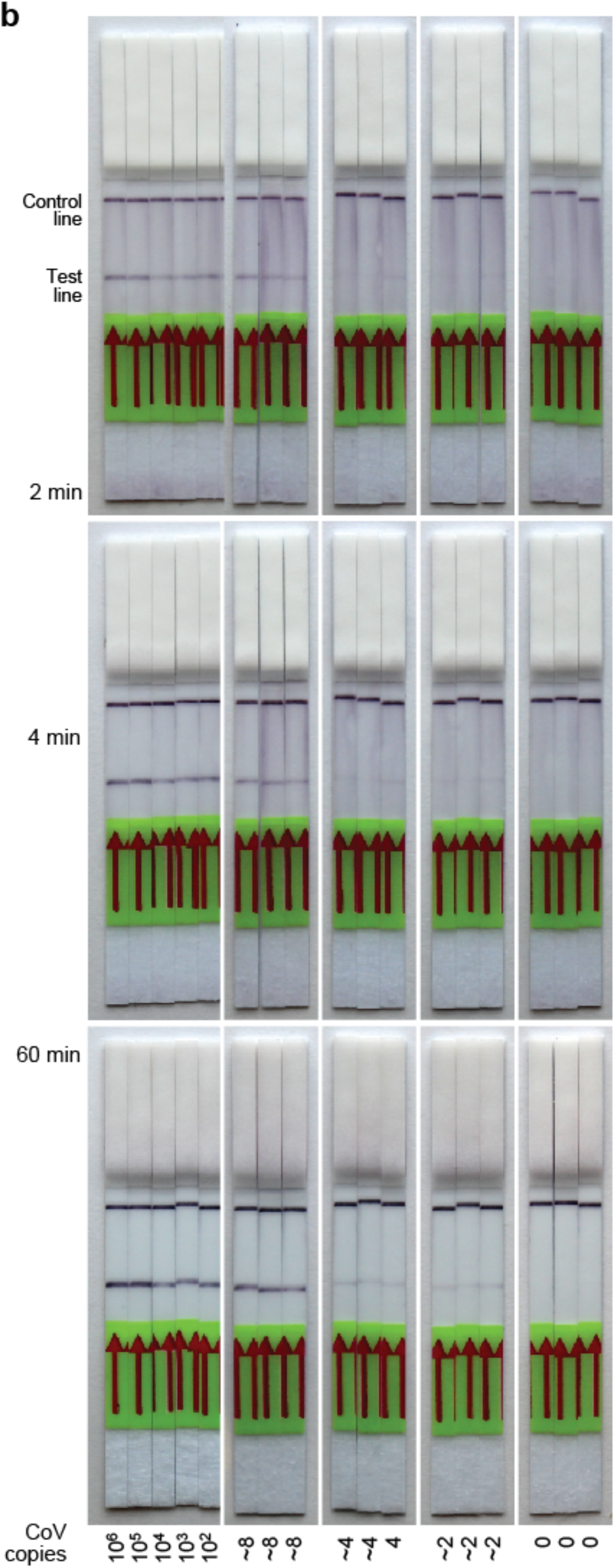
Gels and full-length images of strips in main text Figure 1d. (**a**) A denaturing PAGE gel showing the results of a 5 min RT-RPA for the series dilution of synthetic SARS-CoV-2 RNA shown in main text Figure 1d. Results show strong product bands over a large dynamic range of copy number. (**b**, next page) Full length LFD strips from main text Figure 1d, in which RT-RPA amplicons were subsequently treated via SDS addition, 1 minute T7 exonuclease digestion, and addition of LFD biotin and FAM probes.

**Figure S2:**
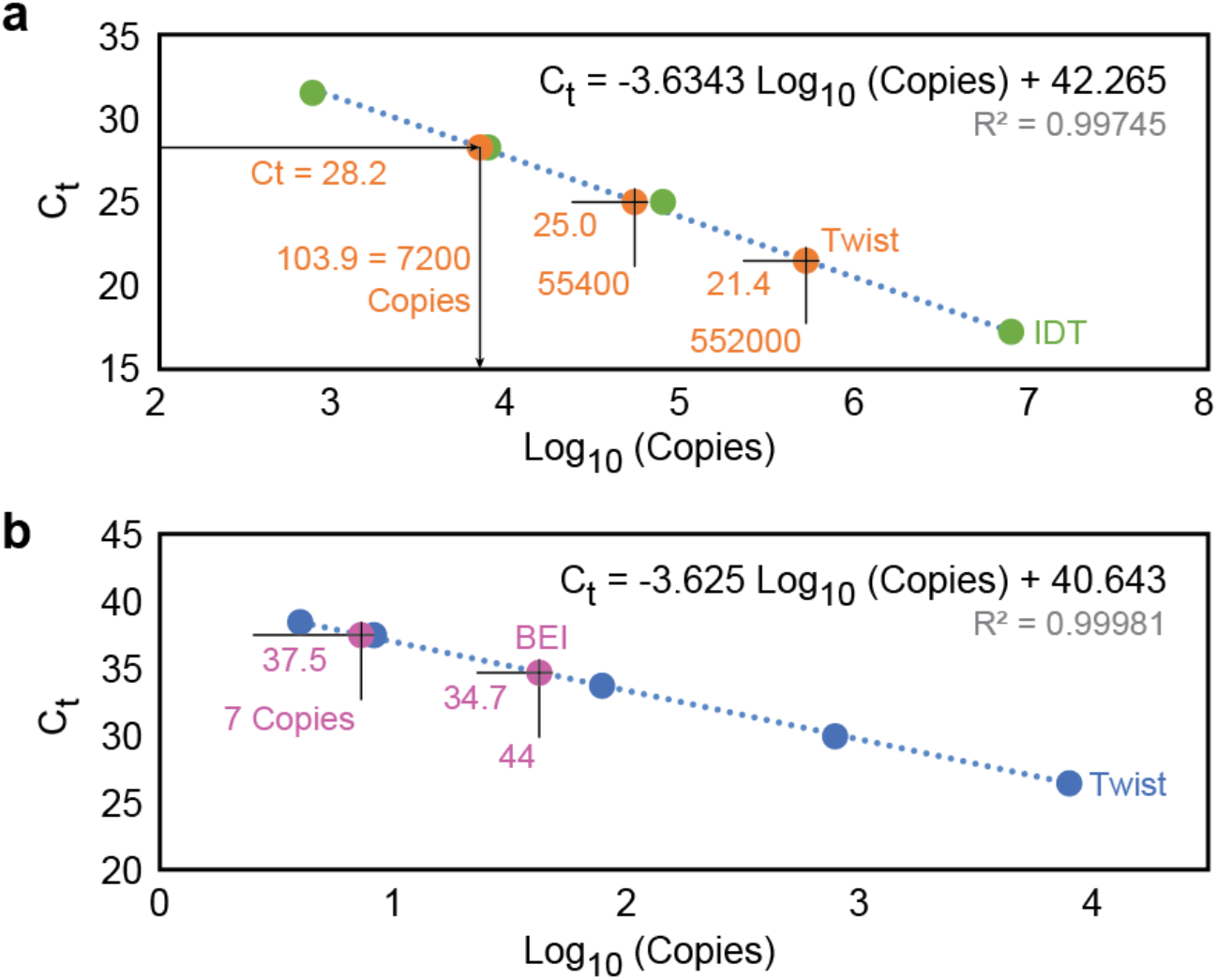
Quantification of spike-in RNA. Synthetic (IDT or Twist) and culture-derived (BEI) SARS-CoV-2 RNA were quantified by spectrophotometry and qPCR for spike-in experiments. (**a**) A 76 nt, 5′ spike sequence RNA segment, synthesized by IDT and shipped at high (∼ 100 µM) concentration, was first quantified by Nanodrop (Thermo Fisher) spectrophotometry. The machine was calibrated and “blanked” with RNA-free buffer, and absorbance at 260 nm (A260) was recorded for IDT RNA in the same buffer in triplicate. The average value (69.3 ±0.15) was used in conjunction with the sequence-specific extinction coefficient (756800 /M/cm) to calculate a stock concentration of 91.6 µM. This RNA was diluted and spiked at ∼ 8E2 to 8E6 copies per 5 µl into RT-qPCR reactions, with our ssRPA 5′ spike primers and EvaGreen dye, and RT-qPCR was performed in triplicate as described in Methods. Values for C_t_ were recorded and a least squares fit was made between C_t_ and known copy number (shown), with R^2^ >0.997. In separate wells of the same run, 3 dilutions of Twist synthetic SARS-CoV-2 RNA were amplified in an identical manner, and those C_t_ values were converted to copy numbers by applying the described fit, yielding 7200 to 552,000 copies per 5 µl. This quantified Twist sample was used in the spike-in buffer experiments of main text Fig. 1d. (**b**) This quantified Twist sample was then used in a dilution series to quantify the culture-derived (BEI) RNA at very low copy, to ensure that low-copy ssRPA spike-in experiments were accurate. As in (a), the Twist and BEI samples were both amplified by RT-qPCR, this time using Charite “E” primer and sequence-specific probe set (IDT) for maximum accuracy at low copy number. The Twist results were fit to the relation shown, in turn used to quantify BEI copy numbers to 44 and 7 per sample, as shown. These BEI dilutions were used in the spike-in saliva and specificity experiments of main text Fig. 1e and 1f.

**Figure S3:**
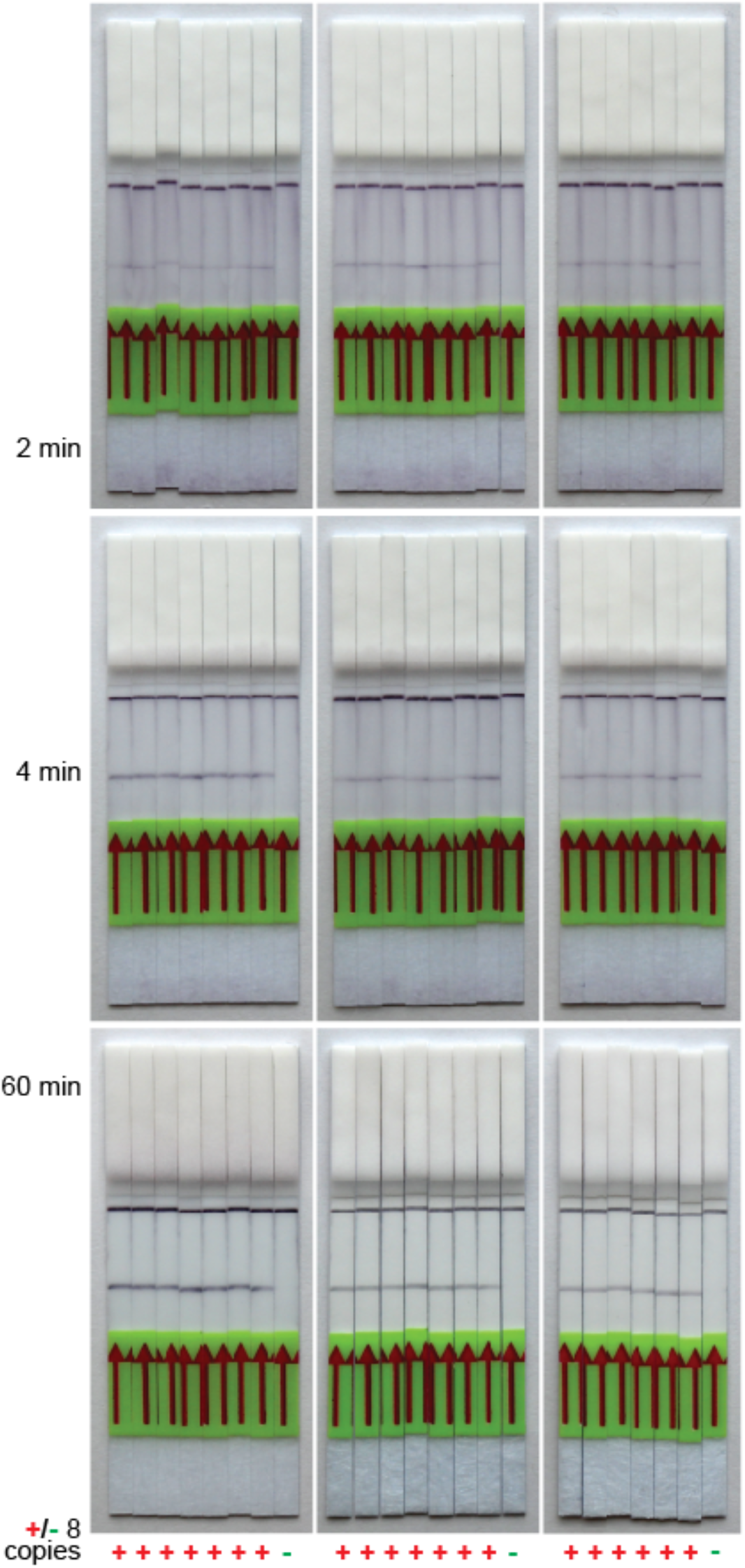
Full-length LFDs of main text Fig. 1e. LFDs were run as in main text, with 0 or 8 copies of cultured, heat inactivated SARS-CoV-2 virus (BEI) per reaction spiked in human pooled saliva, as the LFD incubation times indicated. LFD images were cropped and adjusted for contrast to appear as viewed by the naked eye.

**Figure S4:**
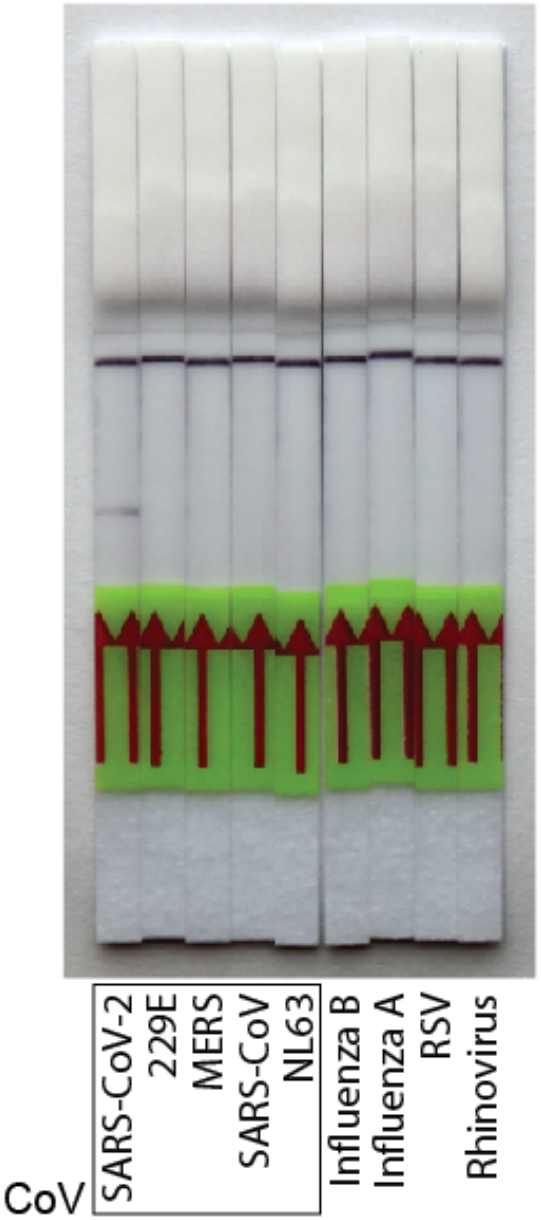
Full-length LFDs of main text Fig. 1f. LFDs were run as in main text, with high copy genomic RNA from the alternative diagnosis viruses shown. LFDs shown at 10 min to indicate lack of false positive bands. LFD images were cropped and adjusted for contrast to appear as viewed by the naked eye.

**Figure S5:**
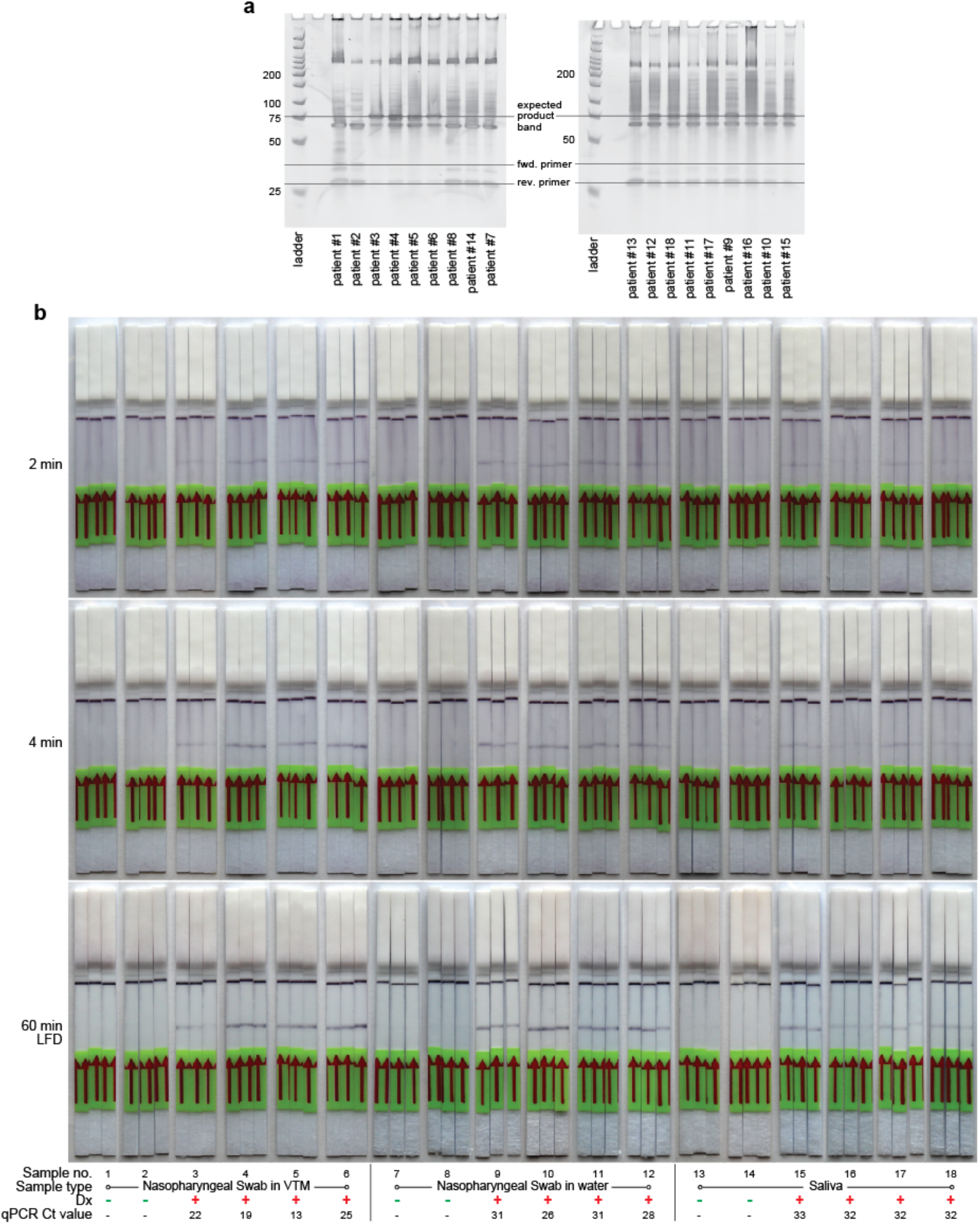
Gel and full-length LFDs of main text Fig. 1g. (**a**) A denaturing PAGE gel showing the results of a 5 min RT-RPA and subsequent SDS addition, 1 min T7 exonuclease digestions, and addition of LFD biotin and FAM probes, for the clinical samples shown in the first of each triplicate of main text Fig. 1g. (**b**) Full length LFD strips from main text Fig. 1g.

**Figure S6:**
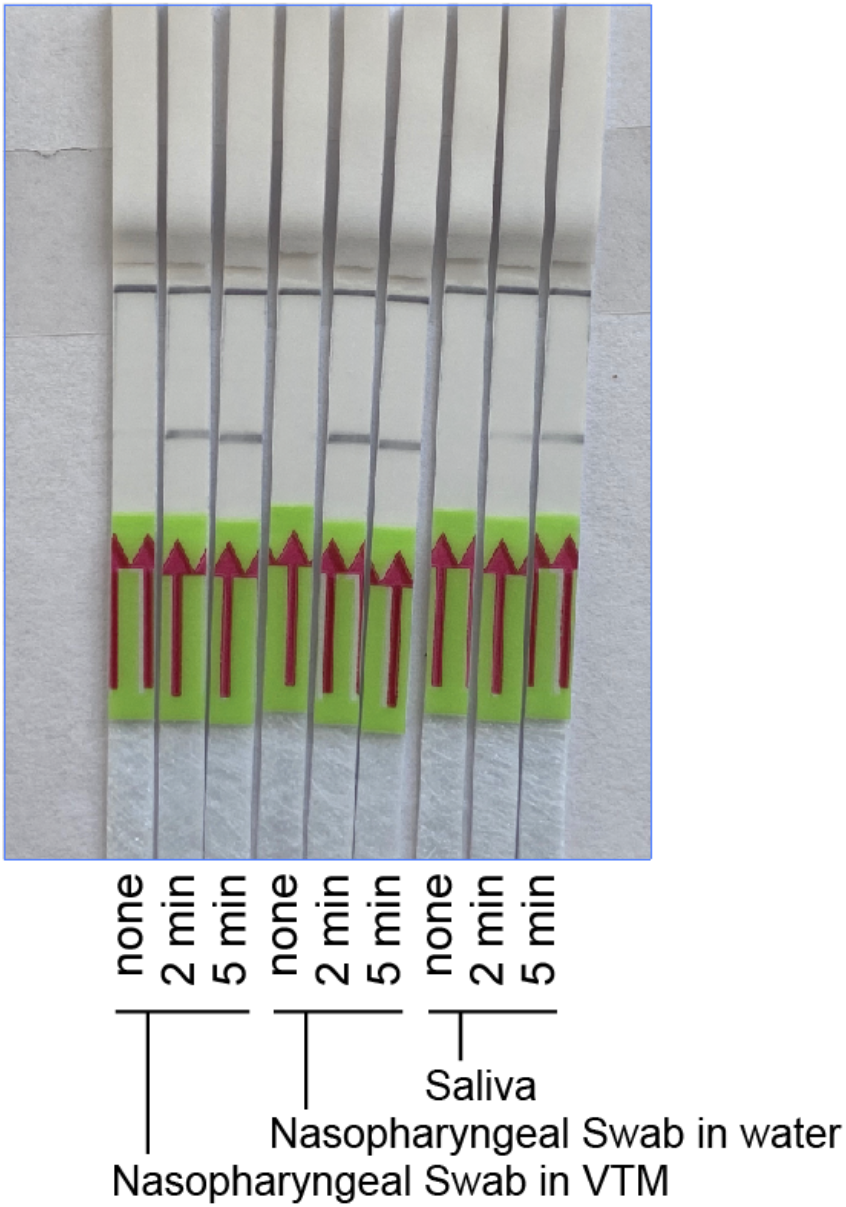
Demonstration of efficacy of extraction protocol. Clinical samples were received from vendor after heat inactivation (95 °C × 5 min) to protect researchers. The ssRPA protocol applied directly to shipped samples resulted in little or no signal, but the 2 min extraction protocol with Lucigen DNA extraction buffer (see Methods) was both necessary and sufficient to yield positive results. Strips were photographed at 60 min.

**Figure S7:**
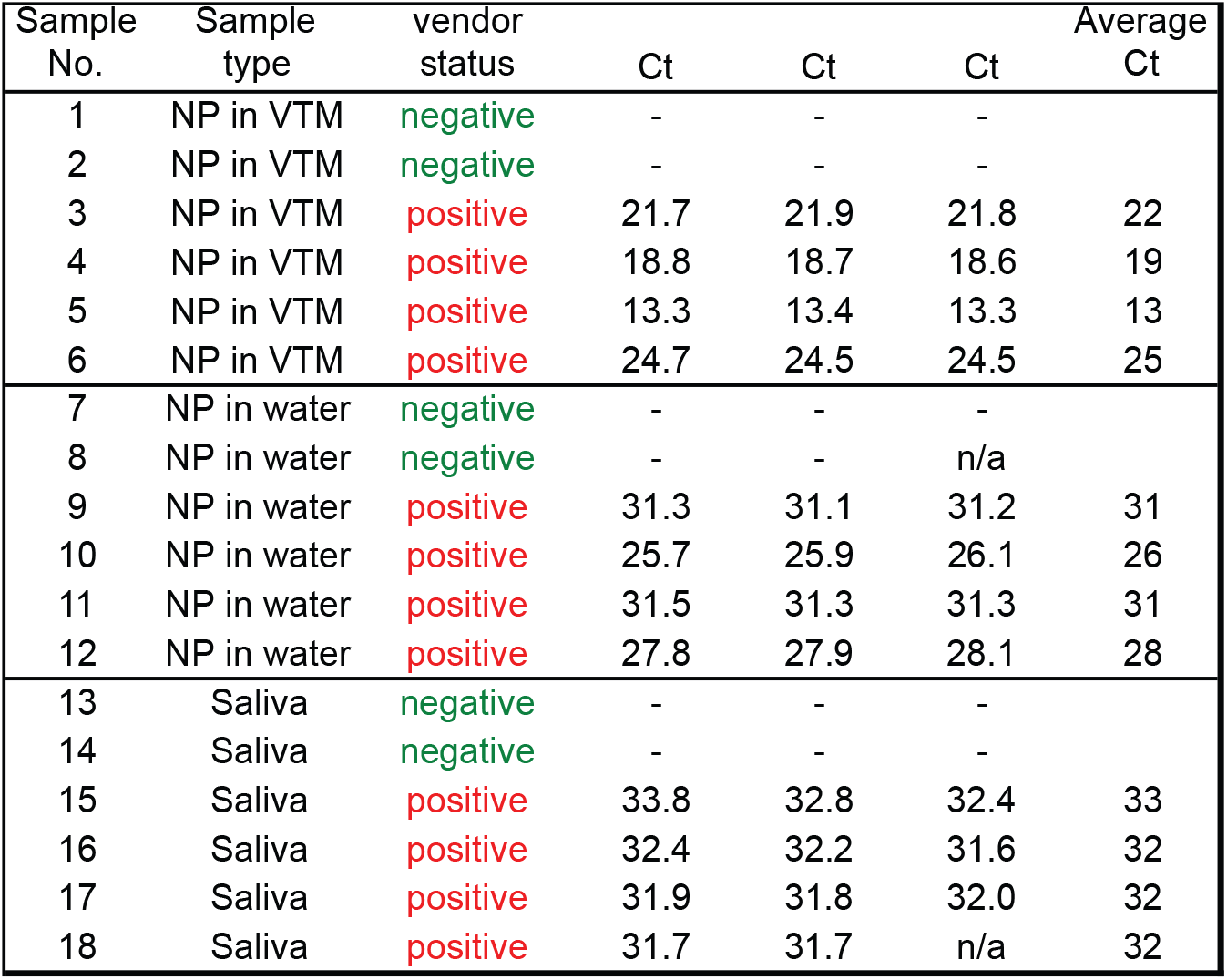
Clinical sample qPCR quantification. Clinical samples were received from vendor with assigned SARS-CoV-2 status based in vendor qPCR. We repeated qPCR quantification in a single experiment to compare relative C_t_ values across samples and to confirm RNA status. Volumes of 100 µl of each raw clinical sample were extracted and buffer exchanged, per CDC qPCR recommendations, with the Qiagen QiaAMP viral RNA mini kit, eluting into 20 µl. Column-extracted volumes of 5 µl each were used in 20 µl qPCR reactions with CDC N2 primers and probes and TaqPath 1-step RT-qPCR master mix, per Methods and CDC recommendations. Extracted samples were tested in triplicate when extraction volumes allowed (“n/a” indicates volumes were insufficient). Forty-four temperature cycles were applied, and those that did not cross the C_t_ threshold after 44 cycles are listed as “-”. Average C_t_ values are also shown and correspond to those in main text Fig. 1g. In other experiments not shown, it appeared that small column elution (extraction) volumes (meant to concentrate samples) led to a 5-fold or higher variation in column elution yield. Quantification C_t_ values shown are therefore only accurate to ∼ +/- 2 cycles. Ranges nevertheless suggest relatively high RNA concentrations for NP swabs in VTM compared to NP swabs in water or in raw saliva.

## References

1. CDC 2019-Novel Coronavirus (2019-nCoV) Real-Time RT-PCR Diagnostic Panel Instructions for Use.pdf.

2. Wee, S. K., Sivalingam, S. P. & Yap, E. P. H. Rapid direct nucleic acid amplification test without RNA extraction for SARS-CoV-2 using a portable PCR thermocycler. http://biorxiv.org/lookup/doi/10.1101/2020.04.17.042366 (2020) doi:10.1101/2020.04.17.042366.

3. Ladha, A., Joung, J., Abudayyeh, O. O., Gootenberg, J. S. & Zhang, F. A 5-min RNA preparation method for COVID-19 detection with RT-qPCR. 6.

4. Bruce, E. A. et al. RT-qPCR Detection of SARS-CoV-2 RNA from patient nasopharyngeal swab using Qiagen RNeasy kits or directly via omission of an RNA extraction step. bioRxiv 2020.03.20.001008 (2020) doi:10.1101/2020.03.20.001008.

5. Arumugam, A. & Wong, S. The Potential Use of Unprocessed Sample for RT-qPCR Detection of COVID-19 without an RNA Extraction Step. bioRxiv 2020.04.06.028811 (2020) doi:10.1101/2020.04.06.028811.

6. Marzinotto, S. et al. A streamlined approach to rapidly detect SARS-CoV-2 infection, avoiding RNA extraction. medRxiv 2020.04.06.20054114 (2020) doi:10.1101/2020.04.06.20054114.

7. Won, J. et al. Development of a Laboratory-safe and Low-cost Detection Protocol for SARS-CoV-2 of the Coronavirus Disease 2019 (COVID-19). Exp. Neurobiol. 29, 107–119 (2020).

8. Zhao, Z. et al. A simple magnetic nanoparticles-based viral RNA extraction method for efficient detection of SARS-CoV-2. http://biorxiv.org/lookup/doi/10.1101/2020.02.22.961268 (2020) doi:10.1101/2020.02.22.961268.

9. Brown, J. R. et al. Comparison of SARS-CoV2 N gene real-time RT-PCR targets and commercially available mastermixes. http://biorxiv.org/lookup/doi/10.1101/2020.04.17.047118 (2020) doi:10.1101/2020.04.17.047118.

10. Xpert Xpress SARS-CoV-2 EUA Instructions for Use, rev D.pdf.

11. Wölfel, R. et al. Virological assessment of hospitalized patients with COVID-2019. Nature (2020) doi:10.1038/s41586-020-2196-x.

12. Chandler-Brown, D., Bueno, A. M., Atay, O. & Tsao, D. S. A Highly Scalable and Rapidly Deployable RNA Extraction-Free COVID-19 Assay by Quantitative Sanger Sequencing. http://biorxiv.org/lookup/doi/10.1101/2020.04.07.029199 (2020) doi:10.1101/2020.04.07.029199.

13. Octant SwabSeq SARS-CoV-2 testing. Notion https://www.notion.so.

14. Illumina CovidSeq Test Instructions for Use (# 1000000123387). 36 (2020).

15. Wang, M. et al. Nanopore Targeted Sequencing for the Accurate and Comprehensive Detection of SARS-CoV-2 and Other Respiratory Viruses. Small 2002169 (2020) doi:10.1002/smll.202002169.

16. Piepenburg, O., Williams, C. H., Stemple, D. L. & Armes, N. A. DNA Detection Using Recombination Proteins. PLOS Biol. 4, e204 (2006).

17. Ding, X., Yin, K., Li, Z. & Liu, C. All-in-One Dual CRISPR-Cas12a (AIOD-CRISPR) Assay: A Case for Rapid, Ultrasensitive and Visual Detection of Novel Coronavirus SARS-CoV-2 and HIV virus. http://biorxiv.org/lookup/doi/10.1101/2020.03.19.998724 (2020) doi:10.1101/2020.03.19.998724.

18. Lucia, C., Federico, P.-B. & Alejandra, G. C. An ultrasensitive, rapid, and portable coronavirus SARS-CoV-2 sequence detection method based on CRISPR-Cas12. http://biorxiv.org/lookup/doi/10.1101/2020.02.29.971127 (2020) doi:10.1101/2020.02.29.971127.

19. Zhang, F., Abudayyeh, O. O. & Gootenberg, J. S. A protocol for detection of COVID-19 using CRISPR diagnostics. 8.

20. Metsky, H. C., Freije, C. A., Kosoko-Thoroddsen, T.-S. F., Sabeti, P. C. & Myhrvold, C. CRISPR-based surveillance for COVID-19 using genomically-comprehensive machine learning design. http://biorxiv.org/lookup/doi/10.1101/2020.02.26.967026 (2020) doi:10.1101/2020.02.26.967026.

21. Hou, T. et al. Development and Evaluation of A CRISPR-based Diagnostic For 2019-novel Coronavirus. http://medrxiv.org/lookup/doi/10.1101/2020.02.22.20025460 (2020) doi:10.1101/2020.02.22.20025460.

22. Qian, J. et al. An enhanced isothermal amplification assay for viral detection. http://biorxiv.org/lookup/doi/10.1101/2020.05.28.118059 (2020) doi:10.1101/2020.05.28.118059.

23. Guo, L. et al. SARS-CoV-2 detection with CRISPR diagnostics. Cell Discov. 6, 34 (2020).

24. Abbott ID-Now SARS-CoV-2 EUA Instructions for Use.

25. El-Tholoth, M., Bau, H. H. & Song, J. A Single and Two-Stage, Closed-Tube, Molecular Test for the 2019 Novel Coronavirus (COVID-19) at Home, Clinic, and Points of Entry. 21.

26. Notomi, T. Loop-mediated isothermal amplification of DNA. Nucleic Acids Res. 28, 63e–663 (2000).

27. Gonzalez-Gonzalez, E. et al. Scaling diagnostics in times of COVID-19: Colorimetric Loop-mediated Isothermal Amplification (LAMP) assisted by a 3D-printed incubator for cost-effective and scalable detection of SARS-CoV-2. medRxiv 2020.04.09.20058651 (2020) doi:10.1101/2020.04.09.20058651.

28. Zhang, Y. et al. Rapid Molecular Detection of SARS-CoV-2 (COVID-19) Virus RNA Using Colorimetric LAMP. http://medrxiv.org/lookup/doi/10.1101/2020.02.26.20028373 (2020) doi:10.1101/2020.02.26.20028373.

29. Park, G.-S. et al. Development of Reverse Transcription Loop-Mediated Isothermal Amplification Assays Targeting Severe Acute Respiratory Syndrome Coronavirus 2 (SARS-CoV-2). J. Mol. Diagn. 22, 729–735 (2020).

30. Yan, C. et al. Rapid and visual detection of 2019 novel coronavirus (SARS-CoV-2) by a reverse transcription loop-mediated isothermal amplification assay. Clin. Microbiol. Infect. 26, 773–779 (2020).

31. Yang, W. et al. Rapid Detection of SARS-CoV-2 Using Reverse transcription RT-LAMP method. http://medrxiv.org/lookup/doi/10.1101/2020.03.02.20030130 (2020) doi:10.1101/2020.03.02.20030130.

32. Yu, L. et al. Rapid colorimetric detection of COVID-19 coronavirus using a reverse tran-scriptional loop-mediated isothermal amplification (RT-LAMP) diagnostic plat-form: iLACO. medRxiv 2020.02.20.20025874 (2020) doi:10.1101/2020.02.20.20025874.

33. Jiang, M. et al. Development and Validation of a Rapid, Single-Step Reverse Transcriptase Loop-Mediated Isothermal Amplification (RT-LAMP) System Potentially to Be Used for Reliable and High-Throughput Screening of COVID-19. Front. Cell. Infect. Microbiol. 10, 331 (2020).

34. Broughton, J. P. et al. CRISPR–Cas12-based detection of SARS-CoV-2. Nat. Biotechnol. (2020) doi:10.1038/s41587-020-0513-4.

35. Joung, J. et al. Point-of-care testing for COVID-19 using SHERLOCK diagnostics. http://medrxiv.org/lookup/doi/10.1101/2020.05.04.20091231 (2020) doi:10.1101/2020.05.04.20091231.

36. Sherlock SARS-CoV-2 EUA, Instructions for Use.

37. Wei, S. et al. Field-deployable, rapid diagnostic testing of saliva samples for SARS-CoV-2. http://medrxiv.org/lookup/doi/10.1101/2020.06.13.20129841 (2020) doi:10.1101/2020.06.13.20129841.

38. Rabe, B. A. & Cepko, C. SARS-CoV-2 Detection Using an Isothermal Amplification Reaction and a Rapid, Inexpensive Protocol for Sample Inactivation and Purification. http://medrxiv.org/lookup/doi/10.1101/2020.04.23.20076877 (2020) doi:10.1101/2020.04.23.20076877.

39. Bhadra, S., Riedel, T. E., Lakhotia, S., Tran, N. D. & Ellington, A. D. High-surety isothermal amplification and detection of SARS-CoV-2, including with crude enzymes. http://biorxiv.org/lookup/doi/10.1101/2020.04.13.039941 (2020) doi:10.1101/2020.04.13.039941.

40. Esbin, M. N. et al. Overcoming the bottleneck to widespread testing: A rapid review of nucleic acid testing approaches for COVID-19 detection. RNA rna.076232.120 (2020) doi:10.1261/rna.076232.120.

41. TwistAmp DNA Amplification Kits: Assay Design Manual.

42. Powell, M. L. et al. New Fpg probe chemistry for direct detection of recombinase polymerase amplification on lateral flow strips. Anal. Biochem. 543, 108–115 (2018).

43. Xia, S. & Chen, X. Ultrasensitive and Whole-Course Encapsulated Field Detection of 2019-nCoV Gene Applying Exponential Amplification from RNA Combined with Chemical Probes. (2020) doi:10.26434/chemrxiv.12012789.v1.

44. Kellner, M. J., Koob, J. G., Gootenberg, J. S., Abudayyeh, O. O. & Zhang, F. SHERLOCK: nucleic acid detection with CRISPR nucleases. Nat. Protoc. 14, 2986–3012 (2019).

45. Han, D. et al. Single-stranded DNA and RNA origami. Science 358, eaao2648 (2017).

46. Sayers, J. R., Schmidt, W. & Eckstein, F. 5′–3′ Exonucleases in phosphorothioate-based oligonucleotide-directed mutagenesis. Nucleic Acids Res. 16, 791–802 (1988).

47. Joung, J. et al. Point-of-care testing for COVID-19 using SHERLOCK diagnostics. medRxiv 2020.05.04.20091231 (2020) doi:10.1101/2020.05.04.20091231.

48. Wahed, A. A. E. et al. Recombinase Polymerase Amplification Assay for Rapid Diagnostics of Dengue Infection. PLOS ONE 10, e0129682 (2015).

49. Yurke, B., Turberfield, A. J., Mills, A. P., Jr., Simmel, F. C. & Neumann, J. L. A DNA-fuelled molecular machine made of DNA. Nature 406, 605–608 (2000).

50. Zhang, D. Y., Chen, S. X. & Yin, P. Optimizing the specificity of nucleic acid hybridization. Nat. Chem. 4, 208–214 (2012).

